# Sampling bias minimization in disease frequency estimates

**DOI:** 10.1101/2021.05.04.21256614

**Authors:** Oshrit Shtossel, Yoram Louzoun

## Abstract

An accurate estimate of the number of infected individuals in any disease is crucial. Current estimates are mainly based on the fraction of positive samples or the total number of positive samples. However, both methods are biased and sensitive to the sampling depth. We here propose an alternative method to use the attributes of each sample to estimate the change in the total number of positive patients in the total population. We present a Bayesian estimator assuming a combination of condition and time-dependent probability of being positive, and mixed implicit-explicit solution for the probability of a person with conditions *i* at time *t* of being positive. We use this estimate to predict the total probability of being positive at a given day *t*.

We show that these estimate results are smooth and not sensitive to the properties of the samples. Moreover, these results are a better predictor of future mortality.

## 1 Introduction

A central tool in the management of epidemics is a precise estimate of the total number of infected individuals. Multiple methods have been proposed for such estimates [1]. The Centers for Disease Control and Prevention (CDC) estimates the daily “percent positive” by dividing the number of positive tests by the total number (positive and negative) of tests for every given day. In some countries, the definition differs from the CDC. They do not compare between the amounts of daily positive tests to the total daily tests. Instead, by using the ID of the subjects, they delete people who have repeated positive tests. They then calculate the number of new positives out of the total number of daily tests or out of new individuals checked that day (positive and negative).

Both the number of daily sample tests and the composition of the population sampled vary daily. The numbers of newly infected individuals reported by the CDC and similar countries do not incorporate the number of tests. As further shown, the total number of positive samples is clearly correlated with the total number of tests (Fig. 1 F). Moreover, even the fraction of positive tests is a problematic measure, since variations in the composition of the tested population directly lead to a change in the fraction of positive samples. Assuming that individuals with a higher probability of having disease related symptoms also have a higher probability of being tested, increasing the number of tests would lead to sampling individuals with less symptoms, as can be seen in the Israeli data (as further detailed in Fig. 1 A-E). This leads to a reduction in the fraction of positives as a function of the sample size. Thus, current estimates are biased by the sampling depth. Here, we propose instead to use the properties of the tested individuals to estimate the fraction of positives while minimizing the effect of the changing amount and sampled populations in tests.

**Figure 1:**
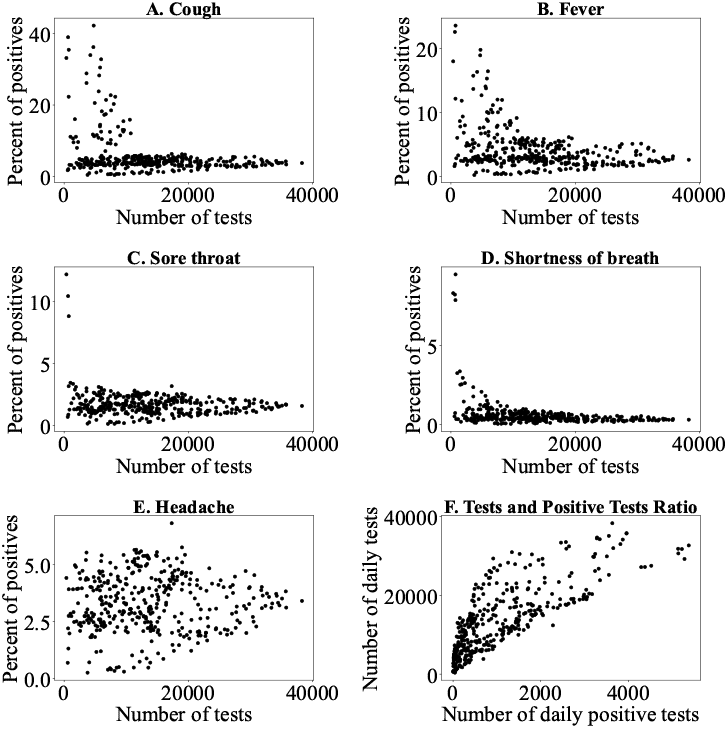
A-E Relation between the number of daily tests and the number of tests being positive for a propriety, based on the Israeli SARS-CoV-2 data between the dates 11.3.2020 and 1.4.2021. In all the properties, except for headache, the fraction of the population positive for the syndrome decreases with the sample size. Subplot F represents the relation between the number of daily tests (x axis) and the number of positive tests (y axis). The daily number of positive tests is highly correlated with the number of daily total tests.

## 2 Related work

Estimates of the total number of infected individuals have been extensively studied [2–4]. Such estimates can be divided into three main tasks:

1. **Estimates of unobserved infected individuals**. Estimates of the total number of infected individuals, based on the observed number of infected individuals [5, 6]. Such methods include, among others, estimate of the failures in infection tracing [7], back estimating infections from mortality [8, 9], and using dynamic statistical techniques [10].
2. **Prediction of future dynamics**. Short or long term prediction of the future number of infected individuals, using epidemic models, fit the observed infection data to different models, often ODE based, to predict future dynamics [11–13]. An interesting model in the current context combines the outer knowledge about COVID-19 with the statistical model of Bayesian analysis trying to find the most likely parameters given the SIR model [14]. A problem with epidemic models is that their results are sensitive to the model parameters, which are often unknown. Future dynamics were also predicted using statistical inference models and time series analysis methods. Such methods address the varying test number problem [15]. They use several statistic parameters, such as the mean, variance, auto correlation, and assuming that these parameters are constant over time [2]. Other statistical methods used are Bayesian inference, Markov Chain Monte Carlo (MCMC) sampling method, etc [16] for a similar goal.
3. **Estimates of the probability of being infected given properties, using machine learning**. Machine learning [17, 18] tools actually use the information of the tested individual and relate it to the probability of having a positive test. However, such tools are not used to estimate the number of infected individuals. Instead, they are used to predict whether a tested individual with given properties is positive.

## 3 Adaptive Bayesian Condition Dependent Estimate -ADVANCE

To address the varying test population composition, the profile of each test should be taken into account. A naïve solution to this problem is to divide the daily sample tests to sub-groups with similar profiles. Let us denote the number of total tests on specific day *t* with a specific profile *i* (a profile is a set of properties - e.g., age, gender and whether the individual has fever) as *N*_*i,t*_ and the parallel number of positive tests as *K*_*i,t*_. The fraction of positive tests at day *t* and a given profile *i* is the ratio of *K*_*i,t*_ and *N*_*i,t*_. If the sample size was large compared with the possible feature combinations, it would be possible to estimate the total probability of being positive, as:

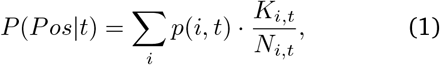

where *p*(*i, t*) is the relative frequency of profile *i* at day *t* in the total population. In reality, the sample size is limited, and the amount of samples in each sub-group is small, and *p*(*i, t*) is not known. We here propose to merge the different groups into a coherent context. We follow here an example using the Israeli SARS-CoV-2 test data.

We merged two datasets. One is the Israeli SARS-CoV-2 data between the dates 11.3.2020 and 1.4.2021. The data was taken from data.gov and is the official data of the Israel Ministry of Health. This data consists of the individual’s features at the Covid-19 test, including: cough, fever, sore throat, shortness of breath, headache, gender, reason for check and age group (60 or above). We used that to estimate *N*_*i,t*_ and *K*_*i,t*_ in Israel [19]. The other is the data of deaths from Covid-19 in Israel per day. It consists of the number of deaths per day in Israel between the dates 11.3.2020 to 9.4.2021. This data was collected from the Israel Ministry of Health Covid dashboard. We used it to validate our models [20].

## 4 Methods and Results

Using the notation above of the number of positive tests on day *t* with specific set of features *i* as *K*_*i,t*_, and the parallel number of tests as *N*_*i,t*_, we further denote *I* as the set of all proprieties combinations, *T* as the set of all days, and *K*_*t*_ = ∑_*i*_*K*_*i,t*_, *N*_*t*_ =∑_*i*_ *N*_*i,t*_ as the total number of positive and overall samples at day *t*, respectively. By using the current estimates of either the total daily number of positive tests *K*_*t*_ or the same value divided by the total daily number of tests 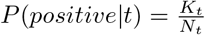, one obtains two different patterns of the epidemics (Figure 2 for the Israeli SARS-CoV-2 data). When using non-normalized values (*K*_*t*_), the patterns are highly sensitive to the number of samples, while in the normalized values 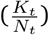, the opposite happens with limited differences in the total positive fraction.

**Figure 2:**
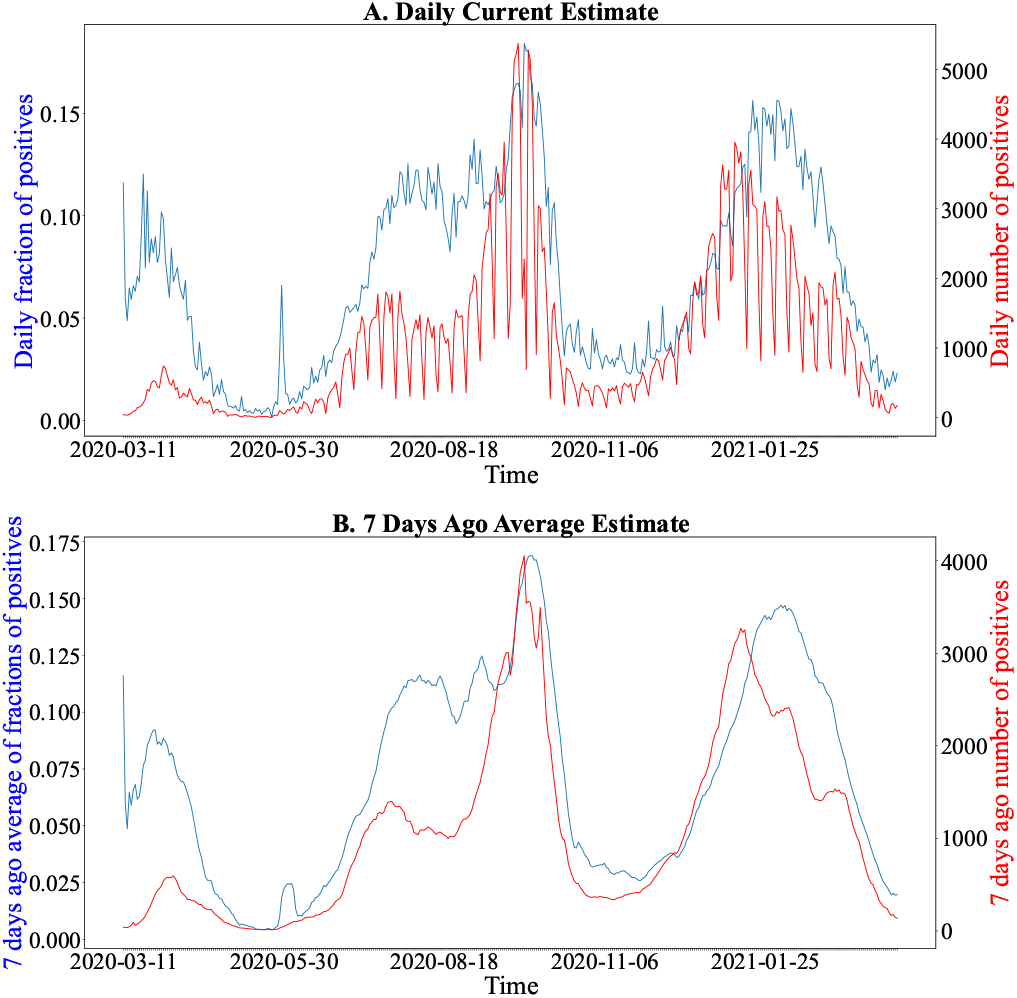
current estimates: X axis - days of tests between 11.3.2020 and 1.4.2021. Y axis - proxies for number of infected. The blue line represents the the fraction of positive tests 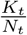. The red line represents the number of positive tests K_t_. The upper plot is the raw data (which has a strong week-end effect). The lower plot is a moving average of 7 days until the current date, except for the first 7 days which do not have 7 previous days.

To reduce the bias in Figure 2, we use a limited set of assumptions:

- The probability of being positive on a certain day with a specific set of features can be approximated by a product of two factors, one that only depends on the date, referred to as *p*_0_(*t*), and the other that depends on the proprieties of the tested individuals and possibly on the date, but is changing very slowly, referred to as *q*_*i*_(*t*). In the initial analysis, we assume *q*_*i*_(*t*) = *q*_*i*_, and then relax this assumption. The probability of being positive on a given day with a given profile is:

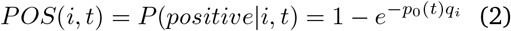
- *p*_0_(*t*) is a smooth function.

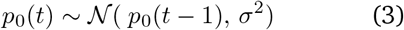 In the following models we chose *σ* to be 0.01, but show that the results are not sensitive to the value of *σ*.
- We consider each sample at day *t* with profile *i* as an independent measure with success probability *POS*(*i, t*) (This may be imprecise if some people tend to do more repeated tests than others, but this does not seem to be a problem in the Israeli data). The resulting distribution of results is binomial with *POS*(*i, t*) as a success probability, with *N*_*t*_ measurements (Eq. 4):

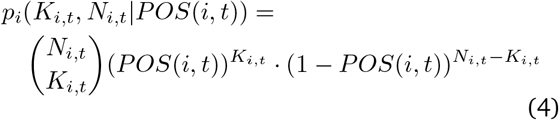

We then estimate *p*_0_(*t*) and *q*_*i*_ [21], by maximizing the log likelihood of:

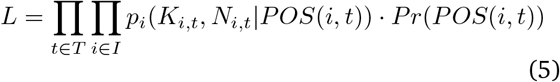

We further assume a uniform beta prior on *q*_*i*_; and a prior for *p*_0_(*t*) as in equation 3, and *Pr*(*POS*(*i, t*)) is a prior on *POS*(*i, t*). We compute ∀*t*; *p*_0_(*t*) and ∀*i*; *q*_*i*_ that maximize the log likelihood. To avoid a large variance in combinations of *i* and *t* with very small sample, we also use a beta prior on the binomial distribution. We tested that the coefficients of the prior are of minimal effect [22]. Specifically, we use the Laplace correction with beta function coefficient of *α* = 1, *β* = 1∀*i*. Leading to:

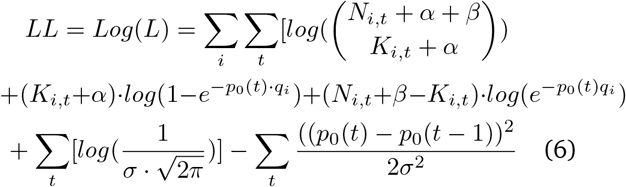

One can now derive *LL* according to *p*_0_(*t*) and *q*_*i*_, and obtain for the *p*_0_(*t*) derivative:

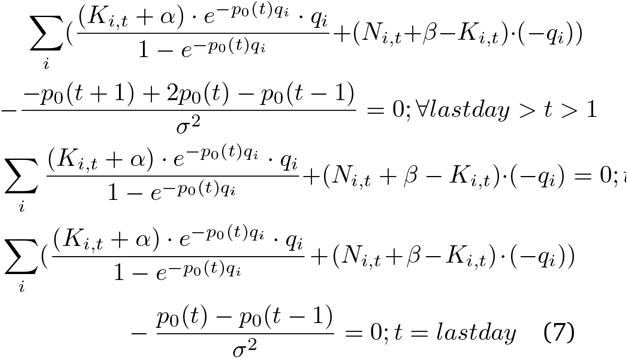

In the *p*_0_(*t*) derivative, we treated each day as if it was the last day, and only computed the back regularization term. Solving with both forward and backward derivative led to numerical instabilities. In the *q*_*i*_ derivative, we approximate 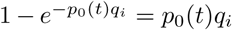, to obtain an explicit solution:

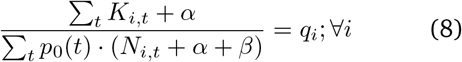

We solve for *q*_*i*_, assuming known *p*_0_(*t*), where *K*_*i,t*_ and *N*_*i,t*_ are observed. We then implicitly solve the *p*_0_(*t*) derivatives numerically. The *p*_0_(*t*) values were found using the scipy Levenberg–Marquardt algorithm [23] as implemented in scipy.root. We initiate the solution by defining the fraction of samples belonging on a given day to a profile *i*:

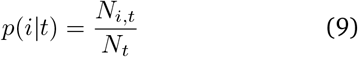

and then initiate the solution for *p*_0_(*t*) using the fraction of positives the same day divided by the average fraction of positives:

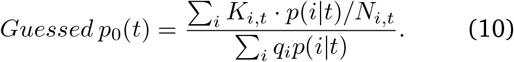

Following the numerical solution of *p*_0_(*t*), we estimate the total fraction of infected individuals to be:

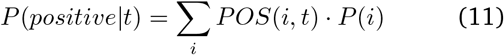

while *P* (*i*) is the total probability of having a specific set of features, i, among the total tests in all sampled days, calculated by:

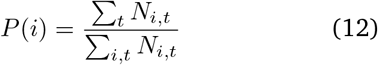

Note that we here assume that while the fraction of samples belonging to a given profile in any given day can be biased, the average over the entire period are representative of the entire population. We have applied the ADVANCE framework on the Israeli data. The results are demonstrated in Figure 3.

**Figure 3:**
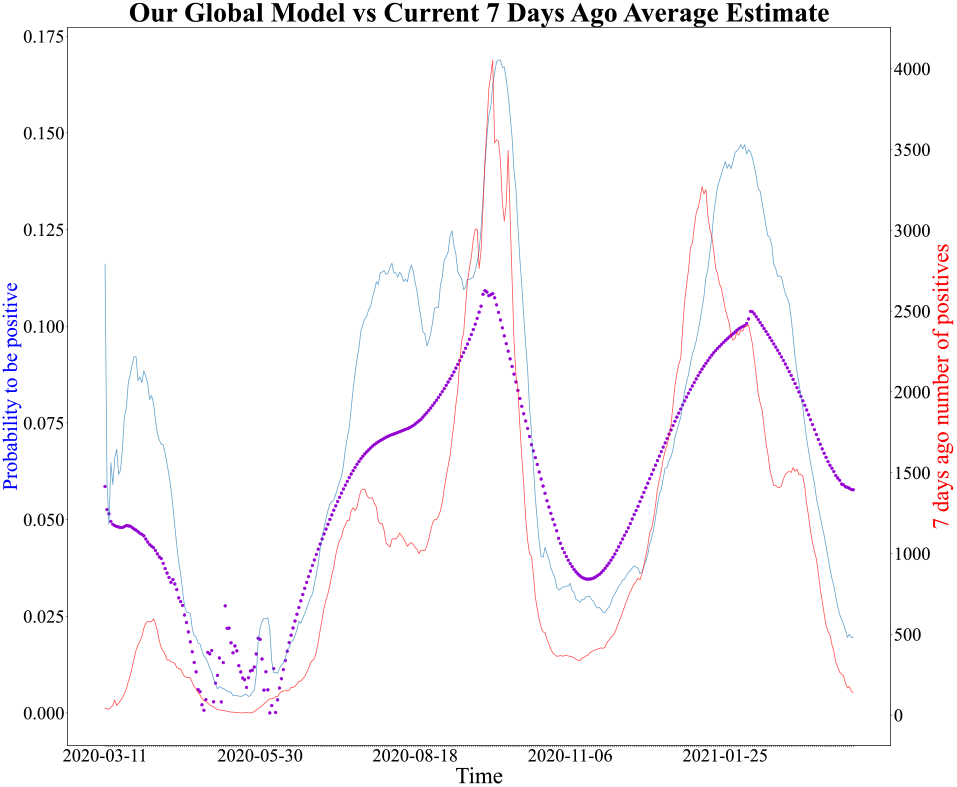
Global vs current estimates: Probability of being positive every day (y axis) during the period 11.3.2020 to 1.4.2021 (x axis). The purple graph represents the ADVANCE estimate, while the red and blue graphs represent the current methods, as in Fig.2.

Two interesting results emerge. First, the obtained estimate is smooth, and not sensitive to the effects of sampling. Second, the result is halfway between the two extreme estimates of the fraction and number of positives. As such, it is less affected by the biases of the two methods. We will further show that this intermediate estimate is more predictive of future death rates than the two existing methods.

*POS*(*i, t*) does not deviate drastically from the fraction of positive tests *K*_*i,t*_*/N*_*i,t*_ (Figure 4), where the size of the scatter points represents the number of tests done. The ADVANCE estimate correlates *K*_*t*_*/N*_*t*_ with correlation of 0.87 and also correlates *K*_*t*_ with correlation of 0.84. There is a small bias that stems from days with a small number of tests, according to the dots size in Fig. 4. Another interesting result in Fig. 4 is that COVID-19 becomes more common as time passes, as the colors of the points transform from blue in the low values to red in the high values. The colors in Fig.4 change from blue in the first dates to red in the late ones according to the colors of the spectrum.

**Figure 4:**
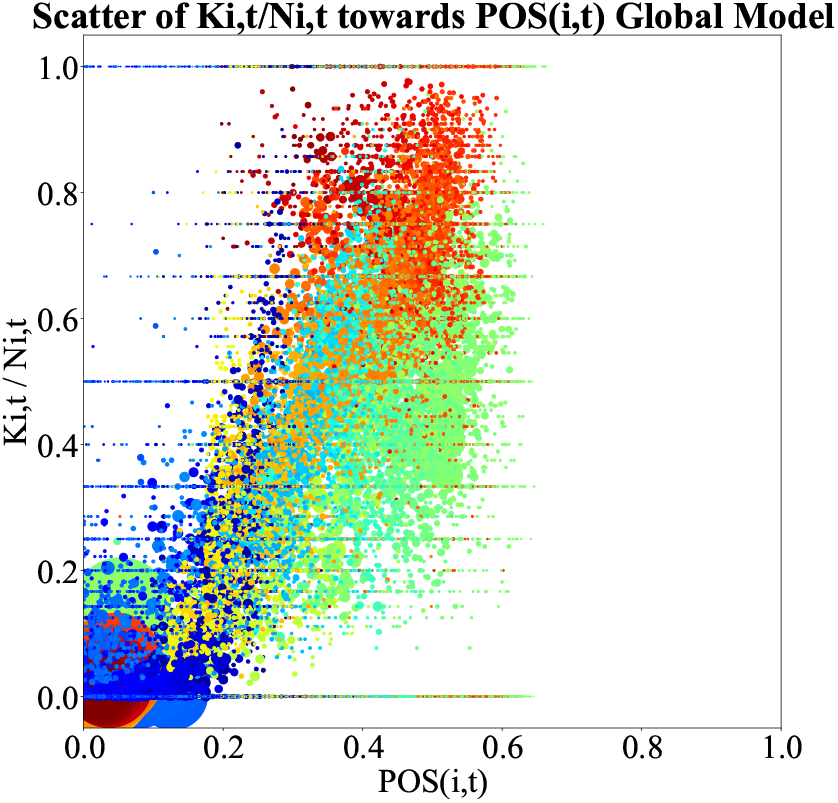
Evaluation: The x axis represents the probability estimated from the bayesian inference, POS(i, t). The y axis represents the probability calculated directly from the data, K_i,t_/N_i,t_. The colors of the points represent the passing days from blue to red, (i.e. probabilities from March are blue and the color of the points that represent the probabilities of later tests changes from blue to red, while the color of the last tests is red). The size of the points represents the number of tests on the day the point represents.The bigger the point is the more tests committed to calculate the probability.

## 5 Online solution

The ADVANCE global is only applicable backward, and is based on a system of equations with an equation and a variable per day. Moreover, it assumes that *q*_*i*_ is static over time. This assumption may be not accurate, over long periods. Thus, the utility of the model above is limited for very long periods. We propose an online version of the same model, with the following assumptions:

- *q*_*i*_ is not static over time. We believe *q*_*i*_ changes slowly within time. We calculate each *q*_*i*_(*t*) according to its 28 previous days, using Eq.8. The first 28 days do not have 28 previous days, therefore their *q*_*i*_(*t*) is constant and equals to the *q*_*i*_ values of the 29th day.
- We find the first 28 *p*_0_(*t*) s by solving the equation system represented in 7 and 8 for only 28 days. In the following days, we find for each consecutive day, *p*_0_(*t*) by solving 7 and 8 assuming *p*_0_(*t*) is known for all days but the last, and only solving the best approximation for the equation root for the last day.
- We update equation 7 in order to deal with the “weekend effect”. The number of tests decreases on the weekend. As a result, assuming that *p*_0_(*t*) is normally distributed around the average of the previous week (and not the day before):

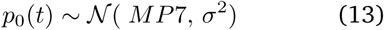

where MP7 is the average of the 7 previous *p*_0_(*t*)s.

Formally, we update equation 7 to be:

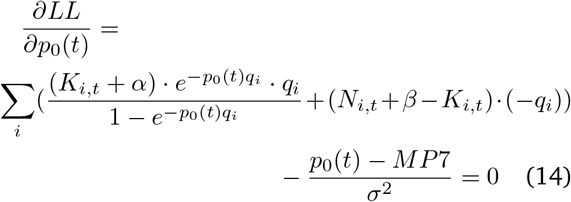

The results of this model are represented in pink in Figure 5, with quite similar results to the global solution.

**Figure 5:**
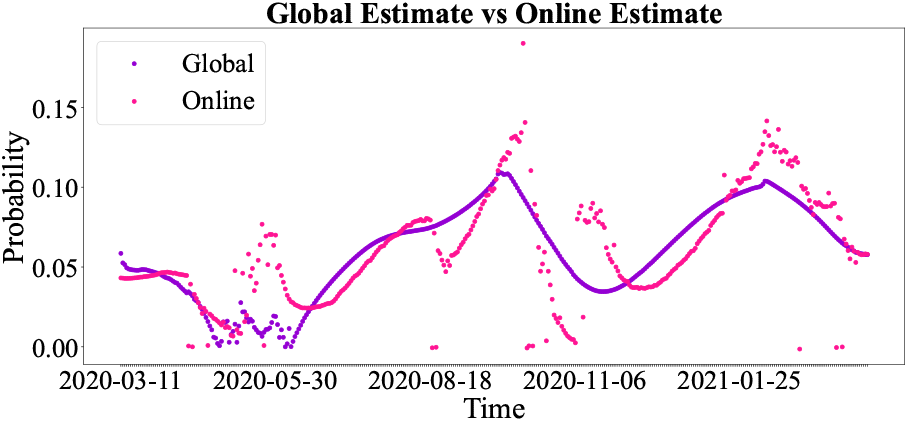
Comparison between global (purple) and online (pink) solutions. The hyper parameters of both models are: alpha, beta equals 1 and sigma equals 0.01. The special hyper parameters for the online model are:number of days to estimate q_i_(t) = 28 and number of days for the p_0_(t) regularization = 7. The models are quite similar. As the time passes they get closer to each other.

## 6 Model Validation

To test the quality of the different estimators, we compared their correlation with future death counts. We used again the Israeli data, and computed the delayed correlation between death events and predicted infectivity. We compared ADVANCE, the fraction of positives and the total number of positives. We used two different values of *σ* for ADVANCE, and used delays of between 0 and 30 days. We analyzed both the global and the online versions. The global model gives by far the best Spearman Correlation (Figure 7) with the number of death events, with an optimal delay of Twenty days. The online model has a similar correlation as the raw number of positive tests, while the positive test fraction has the lowest correlation.

We further compared our results with the seven day smoothed fraction or count of positive tests. Even when smoothing the data with windows of 2-9 days, and using the best smoothing factor for each model and testing for delayed correlations, the same results are obtained and our model provides a better correlation than any other model (Table 1). To test the quality of the different estimators, we compared their correlation with future death counts. We used again the Israeli data, and computed the delayed correlation between death events and predicted infectivity. We compared ADVANCE, the fraction of positives and the total number of positives. We used two different values of *σ* for ADVANCE, and used delays of between 0 and 30 days. We analyzed both the global and the online versions. The global model gives by far the best Spearman Correlation (Figure 7) with the number of death events, with an optimal delay of Twenty days. The online model has a similar correlation as the raw number of positive tests, while the positive test fraction has the lowest correlation.

**Table 1:** Validation after smoothing.

| Model estimate | Window of smoothing | Delay | Spearman correlation coefficient |
| --- | --- | --- | --- |
| Raw fraction | 8 | 15 | 0.8569 |
| Raw number | 9 | 13 | 0.9076 |
| Global sigma = 0.01 | 2 | 20 | 0.9209 |
| Global sigma = 0.1 | 2 | 21 | 0.8892 |
| Online sigma=0.01 | 9 | 8 | 0.8530 |

We further compared our results with the seven day smoothed fraction or count of positive tests. Even when smoothing the data with windows of 2-9 days, and using the best smoothing factor for each model and testing for delayed correlations, the same results are obtained and our model provides a better correlation than any other model (Table 1).

## 7 Sensitivity Analysis

In the current analysis, we ignored the age feature, since in Israel this feature was not checked between the dates 15.4.2020-17.6.2020. We tested the possibility of adding an unknown value for the age category in the period when data was missing. This indeed induced noise, but only during the problematic period of time in samples, as represented clearly in Figure 6. We further tested that the results are not sensitive to any of the hyperparameters in the model. Those include: *α* and *β* from the Beta prior, *σ* from the *p*_0_(*t*) regularization, and the number of days used to compute model in two ways. One is the number of previous days for calculating the dynamic *q*_*i*_(*t*). The other is the number of previous days for the average, which set the expectation of the normal distribution *p*_0_(*t*) is taken from. We checked the impact of changing each hyper-parameter on the model results. We tested the following combinations: *α* equals *β* in range of [0.8,1,1.2], sigma in range of [0.01,0.1,1], number of previous days for dynamic *q*_*i*_ in range of [21,28,35] and number of previous days for *p*_0_(*t*) normal distribution [5,7,9] (Figure. 6).

**Figure 6:**
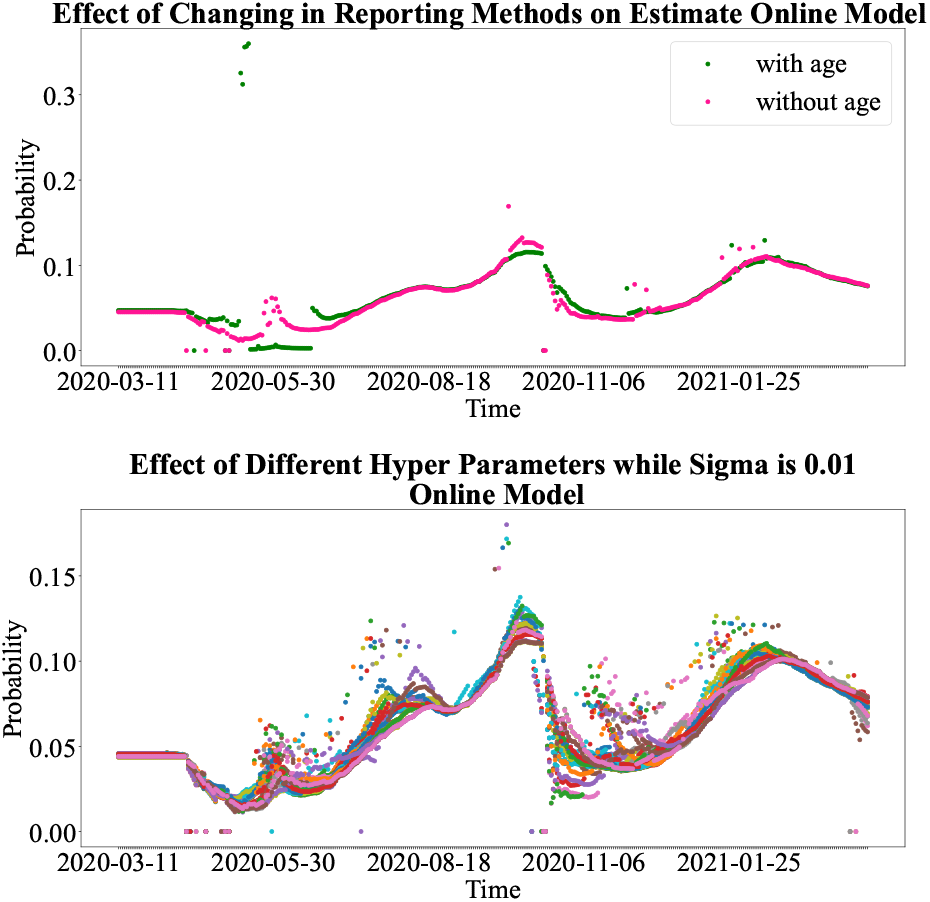
Upper plot Online Analysis - effect of changing input. P(positive|t) (y axis) as a function of time as in previous figures. The Green/Pink points are with/without the age of the tested individual, according to the online estimate. **Lower plot - effect of hyper-variable changes**. Same axes as upper plot. Each color represents another combination of the hyper-parameters of the online model: α and β. See text for parameter range. In both plots, one can see that the variances of the different combinations of hyper-parameters is rather small.

**Figure 7:**
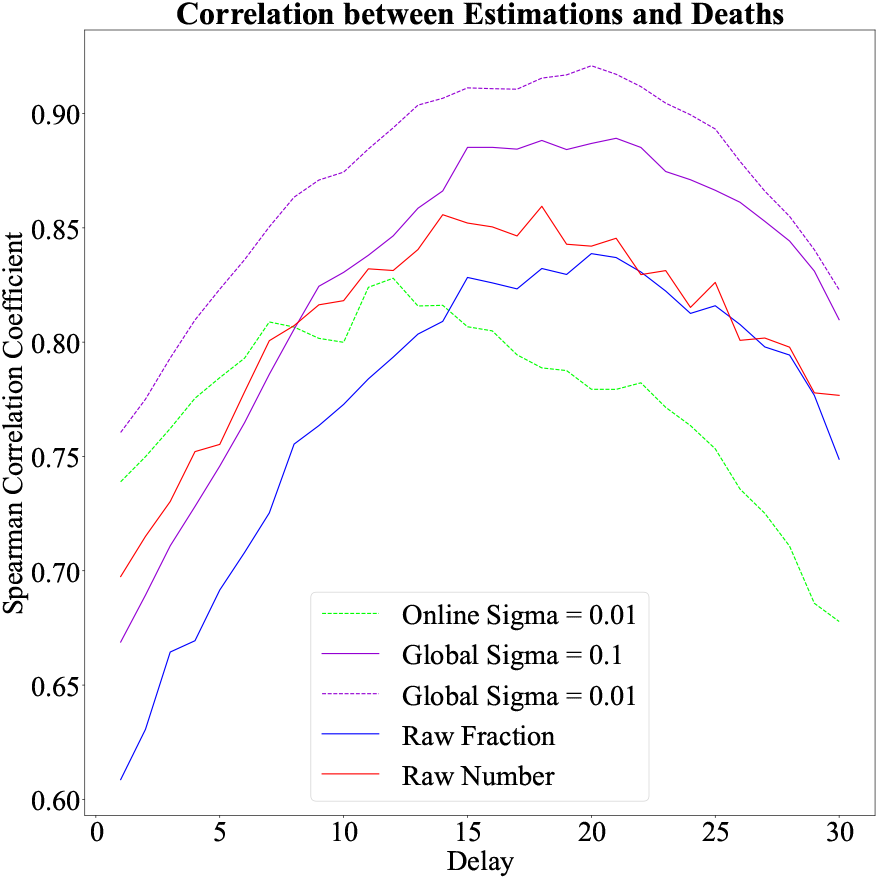
Evaluation: The x axis represents the delay in days between infection and death. The y axis represents the correlation coefficient between the estimate of positive tests and the number of deaths. The global version of ADVANCE gives the best correlations in all the delays we checked.

**Figure 8:**
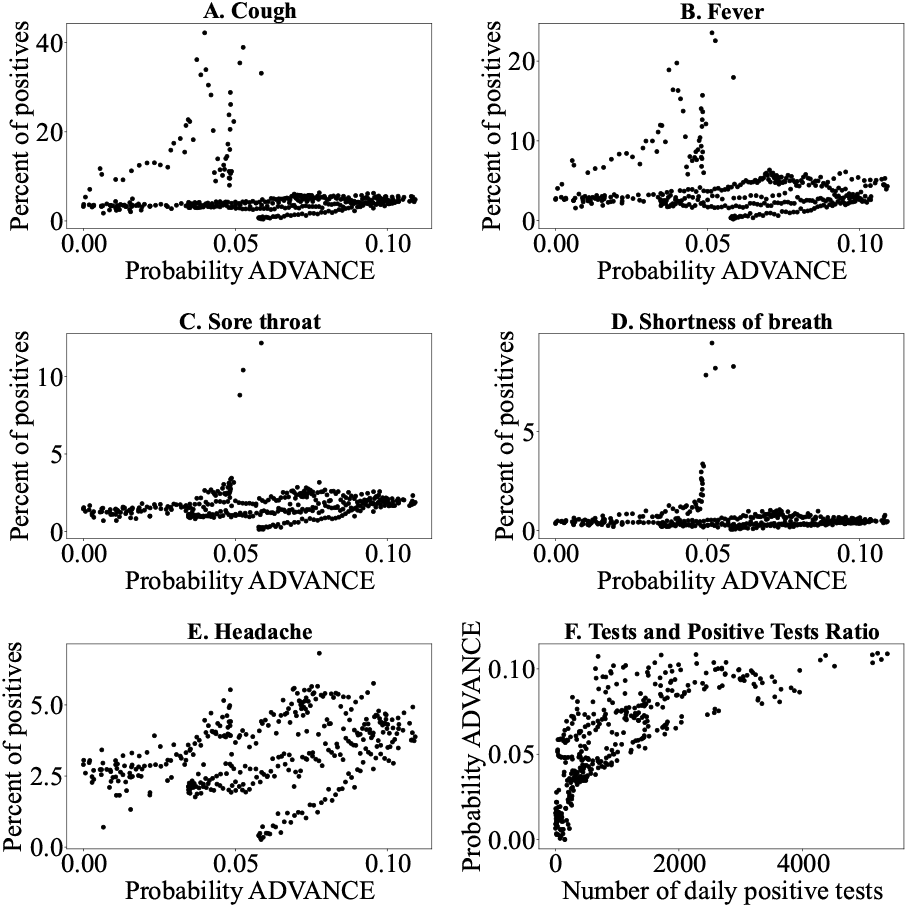
Repetition of the results in Figure 1, but this time not vs the number of tests, but vs the P(positive|t). In contrast with Figure 1, here no correlations are observed.

Similarly, one can change the *β* function priors and the *σ* of the normal distribution and the values of *q*_*i*_ does not change a lot. The different vectors of the *q*_*i*_ values stay highly correlated -Spearman correlation of 0.98.

## 8 Discussion

We suggested two different estimation models to the fraction of daily positives to a certain disease. We used the models on the data of the Israeli Covid-19. On this data, the ADVANCE estimations are much smoother and are less effected by bias than current estimators. Moreover, they are a better prediction of future morbidity. suggested two different estimation models to the fraction of daily positives to a certain disease. We used the models on the data of the Israeli Covid-19. On this data, the ADVANCE estimations are much smoother and are less effected by bias than current estimators. Moreover, they are a better prediction of future morbidity.

The main limitation of the current model is the estimate of *P*(*i*). Even if one knows the property distribution within the sampled population, this distribution may differ from the one in the general population. Moreover, we have here assumed that *P*(*i*) is fixed over time. This may not be the case if, for example, seasonal diseases affect the frequency of people with fever in the population. Still, even with these limitations, ADVANCE is a better predictor of future morbidity than current methods.

Individuals rely on COVID-19 test results to guide their medical treatment and decisions on whether to self-isolate, and to vaccinate. Public health officials rely on the results to track the state of the pandemic, and policymakers use this information to make decisions on reopening schools and businesses. An important factor in these decisions is a proper estimate of the infected population. We thus suggest ADVANCE to improve the quality of such an estimate.

Two versions of ADVANCE were presented - a global aposteriori version and an online version. The global version is a better predictor of future morbidity, but it does not supply a continuous estimate. Running the global version every day with the new data did not produce a smooth enough estimator of the fraction of infected individuals. Both global and online methods include an inherent smoothing of the fraction of infected individuals through the prior on *p*_0_(*t*). As such they can be directly compared to the moving averages of current estimators.

We have here used a set of features defined by a standard questionnaire used in Israel. Most components, except for age, of this questionnaire were found to be informative. However composite indices may end up being more informative. The next stage of the current analysis will be to develop outcome prediction algorithms and through those develop composite features. Another important required extension will be to compare this method in other countries. However, currently, we did not find similar datasets.

## 9 Data Availability

All code and data summaries used in these analyses are posted on https://github.com/oshritshtossel/ADVANCE-Sampling-bias-minimization-in-disease-frequency-estimate

## Data Availability

https://data.gov.il/dataset/covid-19/resource/d337959a-020a-4ed3-84f7-fca182292308
https://datadashboard.health.gov.il/COVID-19/general

## 10 Acknowledgment

We declare no conflict of interests. Funding for this research was by MAFAT grant for Covid dynamics analysis. OS and YL were funded by ISF grant 870/20.

